# Toward a National Registry for Inborn Errors of Immunity in Peru: A Qualitative Implementation Study

**DOI:** 10.64898/2026.06.12.26355539

**Authors:** Liz Veramendi-Espinoza, Kelly De la Cruz-Torralva, Armando Pezo-Pezo, Javier Vargas Herrera, Joan Neyra Quijandria, Martha Martina, Kathleen E. Sullivan, Moises A. Huaman, Jacqueline M. Knapke

## Abstract

**Background:** Peru lacks an integrated information system for patients with Inborn Errors of Immunity (IEI). Although disease registries are essential tools for data management and health planning, their success depends on implementation science approaches that account for local contextual factors. This study reports Phase I of a three-phase mixed-methods implementation project to design and develop a national IEI registry.

**Methods:** Phase I consisted of a phenomenological qualitative study exploring stakeholder perspectives. Semi-structured focus groups and in-depth interviews were conducted with 29 key stakeholders across four groups: policy-makers, clinical experts, end-users (immunologists, residents, allied health personnel), and patient organization representatives. Interviews followed a guide structured around four a priori domains (structure, navigation, feasibility, and perception of existing systems). Discussions were conducted in Spanish, audio-recorded, transcribed verbatim, and coded using ATLAS.ti. A hybrid thematic analysis combining deductive and inductive coding was performed. Data elements proposed for the registry were triangulated with qualitative findings.

**Results:** Thirty-six initial codes were consolidated into 15 categories, which were further integrated into four overarching themes conceptualized as pathways toward intention to use: (1) Environment, where governance, regulatory backing, and sustainable financing were identified as key enablers, while limited interoperability emerged as a structural barrier; (2) Technical Dimension, emphasizing usability, alignment with clinical workflow, and a hierarchical data architecture (demographic–clinical–therapeutic); (3) Users, highlighting clinical leadership, protected time, digital readiness, and perceived usefulness as stronger motivators than financial incentives; and (4) Patients, underscoring data protection, transparency, trust, and advocacy as essential for legitimacy and sustainability.

**Conclusions:** A national IEI registry in Peru is perceived as necessary and feasible if implemented with strong regulatory foundations, interoperable design, robust data security, and user-centered architecture. These findings informed the development of an initial functional prototype and the operational plan for Phase II, focused on usability evaluation.

## Introduction

Inborn errors of immunity (IEI) comprise a heterogeneous group of rare, genetically determined disorders associated with recurrent infections, immune dysregulation, and substantial morbidity(1,2). Delayed diagnosis has been associated with poorer health-related quality of life and higher mortality, whereas early diagnosis may reduce healthcare utilization and generate cost savings ranging from US$6,500 to US$108,463 per patient per year (3,4). Despite growing recognition in Latin America, national epidemiologic and longitudinal follow-up data in Peru remain fragmented, limiting timely diagnosis, coordinated care, and equitable access to therapies(5,6).

The Peruvian health system is highly segmented across multiple public and social security subsystems, with heterogeneous digital maturity, limited interoperability, and uneven specialist distribution concentrated in Lima(7,8). These structural characteristics increase the vulnerability of health information initiatives to fragmentation, duplication, or early discontinuation if governance, technical design, and stakeholder engagement are not carefully aligned(8,9).

IEI-specific registries, such as the European Society for Immunodeficiencies (ESID) Registry, the United States Immunodeficiency Network (USIDNET) Registry, and the Canadian Inborn Errors of Immunity National Registry (CIEINR), help address these gaps by standardizing data capture, enabling routine quality monitoring, and generating real-world evidence for clinical and policy decision-making(10–13). More recent interoperability-oriented initiatives, such as RareLink, further illustrate how modern registry infrastructures can support data harmonization across settings (12,13). However, direct replication of these established registry models may not be feasible or sustainable in low- and middle-income settings such as Peru(9,14). Given the multiplicity of actors involved in IEI care, a qualitative approach was essential to capture divergent priorities, perceived barriers, and shared expectations(15,16). Understanding these perspectives was considered a prerequisite for developing a context-sensitive implementation strategy aligned with technical feasibility, stakeholder needs, and local governance realities(14,17).

This study was structured within the Exploration, Preparation, Implementation, and Sustainment (EPIS) framework(18). The findings reported here correspond to Phase I (Exploration) of a three-phase implementation study aimed at developing Peru’s first national registry for IEI(19). This phase aimed to elicit requirements and priorities from decision-makers, IEI experts, clinical users, and patient-advocacy leaders, and to translate these findings into initial specifications for a context-adapted IEI registry. These outputs were intended to inform the subsequent usability and feasibility phases of the implementation study.

## Materials and Methods

### Study design and setting

We conducted a qualitative phenomenological study as part of the Exploration phase of the EPIS framework(18). A phenomenological approach was selected to explore stakeholders’ experiences, perceptions, and meanings regarding the potential implementation of a national IEI registry in Peru(20). Given the complexity of the health system and the diversity of actors involved, this design allowed in-depth examination of how participants understood feasibility, value, governance, and anticipated use.

Phase I combined semi-structured in-depth interviews and focus groups to elicit requirements for registry content, workflows, governance structures, and implementation conditions(15). Findings from data collection and preliminary analysis were translated through iterative co-design activities into concrete design outputs, including registry workflows, data specifications, and the initial Research Electronic Data Capture (REDCap) prototype for Phase II.

### Participants and sampling

Participants were selected using maximum-variation purposive sampling to ensure heterogeneity in roles, decision-making levels, clinical expertise, and patient representation (21,22). Four stakeholder groups were predefined: (1) decision-makers in health services and policy; (2) national and international IEI experts; (3) clinical users (immunologists, residents, nurses, and allied health professionals working in Clinical Immunology services); and (4) leaders of patient associations representing individuals with IEI or rare diseases.

Stakeholders were identified through a structured mapping process based on formal institutional roles, documented involvement in IEI or rare-disease programs, leadership positions in scientific societies, and recognized participation in patient organizations (Supplementary Table 2). Interviews continued until thematic saturation was achieved across stakeholder groups(23).

### Interview guide and data collection

The interviews and focus groups used semi⍰structured guides that were developed and reviewed by six content experts, two of whom were part of the research team. Questions were organized around four axes derived from the literature, existing registry models, and the study objectives: feasibility (resources, governance, sustainability); data structure (standards, fields, quality); navigation and usability (workflows, interface needs); and perceptions of existing registries (lessons learned from previous systems)(11,24,25).

All four stakeholder groups discussed the four thematic axes. Additionally, Groups II and III (IEI experts and clinical users) participated in a structured “variable card” prioritization exercise. The 78 candidate data items were derived from two sources: core variables from established IEI registries and preliminary consultations within the research team based on national clinical practice(5,10,11). The variable card exercise was used to complement the qualitative discussions. Each item was presented individually and rated on a 5-point Likert scale (1 = not relevant; 5 = essential) to assess its perceived importance for inclusion in a context-adapted minimum data set (MDS) (26,27).

Sessions were conducted using virtual or in-person modalities depending on participant availability. All sessions were conducted in Spanish by two team members: the principal investigator and a psychologist trained in qualitative methods. Sessions were accompanied by field notes, audio-recorded with participant consent when they were in person, and video-recorded with participant consent when they were virtual. Sessions typically lasted approximately 45 minutes, with a small number extending to 90 minutes.

### Data management, transcription, and anonymization

Audio and video recordings were stored on encrypted drives within the secure network of the Telehealth Unit at Universidad Nacional Mayor de San Marcos (UNMSM). Transcripts were generated using automated speech recognition software and translated into English. Subsequently, they were reviewed and manually verified line by line by two members of the research team to ensure accuracy, and supervised by a senior researcher from the University of Cincinnati, who served as an external auditor. All direct identifiers, including personal names, institutional affiliations, and specific locations, were removed or generalized during transcription. Each participant was assigned a unique study identification code. De-identified transcripts were imported into ATLAS.ti for coding and analysis.

### Analytic approach and integration of frameworks

We used a hybrid (inductive–deductive) thematic analysis (28). Two coders performed open coding on an initial subset, meeting to compare interpretations, discuss discrepancies, and generate an initial inductive codebook. Disagreements were resolved through discussion and iterative refinement of code definitions until consensus was reached. After two iterative cycles, we introduced deductive codes to support interpretation: (i) selected EPIS domains to map context and implementation supports, and later (ii) Technology Acceptance Model (TAM) constructs (perceived usefulness/ease of use, attitude, intention) to interpret user-acceptance themes(29). The result was a hybrid codebook that retained emergent codes alongside EPIS/TAM-aligned categories. The harmonized codebook was then applied to the full corpus. We generated code reports and matrices for constant comparison across stakeholder groups and to identify cross-cutting themes.

This qualitative study was reported in accordance with the Consolidated Criteria for Reporting Qualitative Research (COREQ) checklist (30). Details on the research team, study design, data collection, analysis, and reporting are provided in Supplementary Table S1. The research team had prolonged engagement with the data, had regular meetings to discuss the coding of transcripts, and engaged in peer debriefing, where one member of the research team who was familiar with the project but did not participate in the primary coding process acted as a quality and reliability auditor to ensure rigorous thematic analysis.

To triangulate the evidence-based practice (EBP) characteristics, we used a convergent mixed-methods approach that integrated qualitative findings with quantitative prioritization. Qualitatively, EBP characteristics that emerged from interviews and focus groups were classified as either suggested or discouraged. Quantitatively, the Likert-scale “variable card” results were categorized using predefined agreement bands based on the proportion of respondents rating each item as 4 or 5. Items with ≥75% agreement were classified as accepted, consistent with a threshold previously used in registry development studies employing Delphi consensus (27). Items with 50–74% agreement were retained for review, whereas those with <50% agreement were classified as rejected. In our study, these categories were used pragmatically to support prioritization and triangulation rather than as part of a formal Delphi process. Integration of qualitative and quantitative classifications informed the final selection and staging of EBP characteristics, as detailed in Supplementary Table S3.

### Co-design and initial REDCap build

Analytic themes and the item-rating results jointly informed co-design: (1) requirements synthesis into user stories and end-to-end workflows; (2) definition of the MDS using the 78-item ratings to prioritise fields, cross-checked against qualitative themes and standard clinical vocabularies (e.g., the International Classification of Diseases, 10th Revision (ICD-10); the International Union of Immunological Societies (IUIS) classification of inborn errors of immunity); (3) production of wireframes and a data dictionary (field types, branching logic, required fields, validation rules); and (4) configuration of REDCap instruments and role-based permissions. Prototype Version 1 was packaged for Phase II usability testing; detailed design choices and usability outcomes will be reported separately.

### Ethics

The UNMSM Research Ethics Committee approved the protocol (Ref. FMH-UNMSM/CEI-2025-022). All participants provided written informed consent, including permission to record and to use anonymized quotations. Data handling complied with the Peruvian Personal Data Protection Law (Ley N.° 29733). The data obtained in this study (including audio recordings, transcripts, and digital documents) are securely maintained by the principal investigator. They will not be disclosed, except in response to a legal requirement duly authorized by the ethics committee that approved the study.

### Artificial Intelligence Tools and Technologies

Generative artificial intelligence tools (ChatGPT, OpenAI) were used to assist with English-language refinement during manuscript preparation. The authors reviewed, edited, and validated all AI-assisted outputs and retained full responsibility for the accuracy, interpretation, and final wording of the manuscript. No primary study data, transcripts, quotations, coding, analyses, results, or conclusions were generated by AI tools.

## Results

A total of 29 stakeholders participated, representing decision-makers, IEI experts, clinical users, and patient organizations across multiple subsystems of the Peruvian health system. The diversity of roles and institutional positions enabled the examination of structural, technical, professional, and patient-centered dimensions that shape the potential implementation of a national IEI registry. An overview of stakeholder characteristics and institutional affiliations is provided in Supplementary Table S1. Across the interviews and focus groups, an initial set of 36 analytic codes was identified through hybrid thematic analysis. Through iterative comparison and conceptual refinement, these were consolidated into 15 higher-order categories to reduce analytic fragmentation and enhance interpretability. The complete coding structure is summarized in Table 1.

**Table 1.**
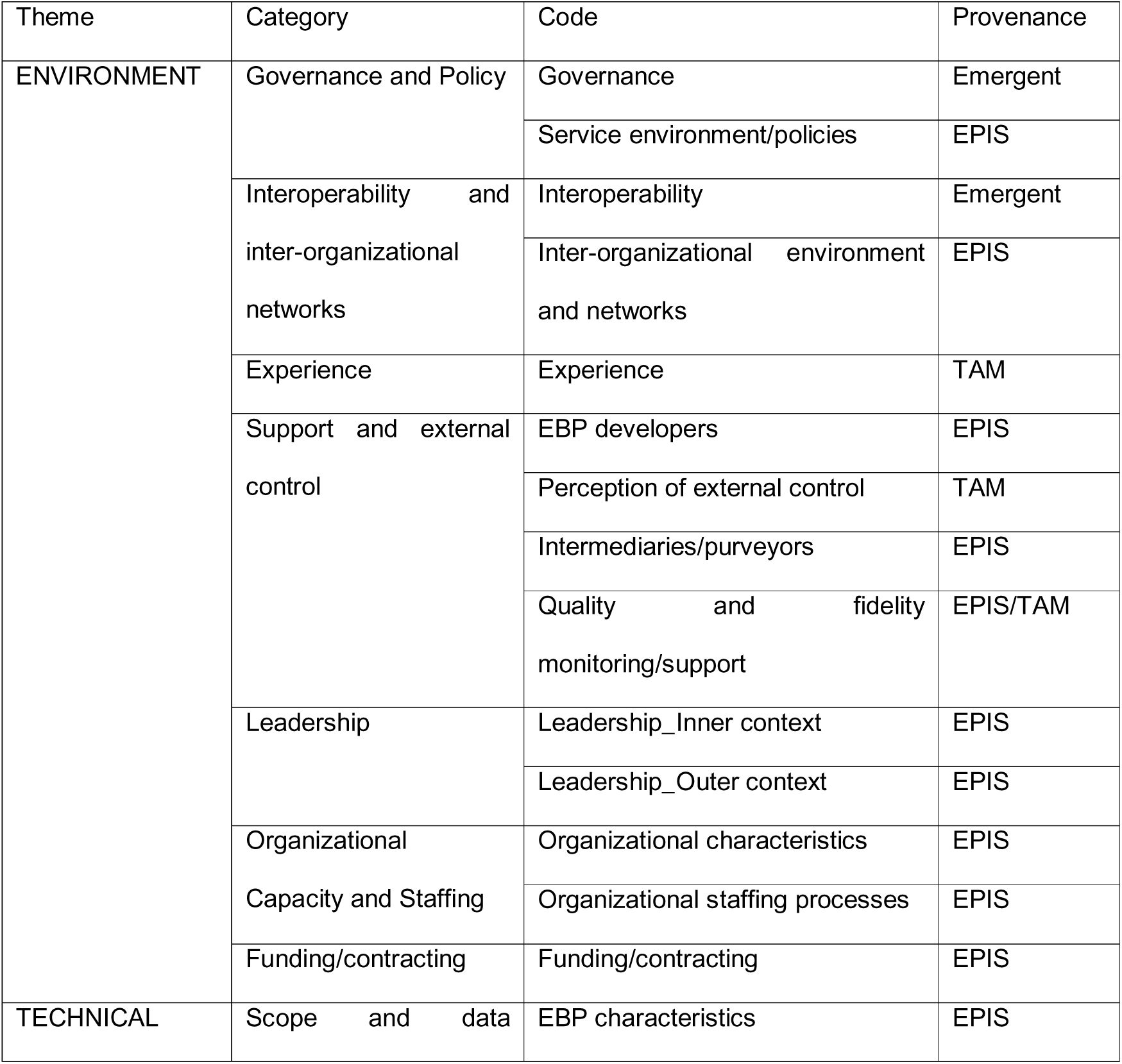
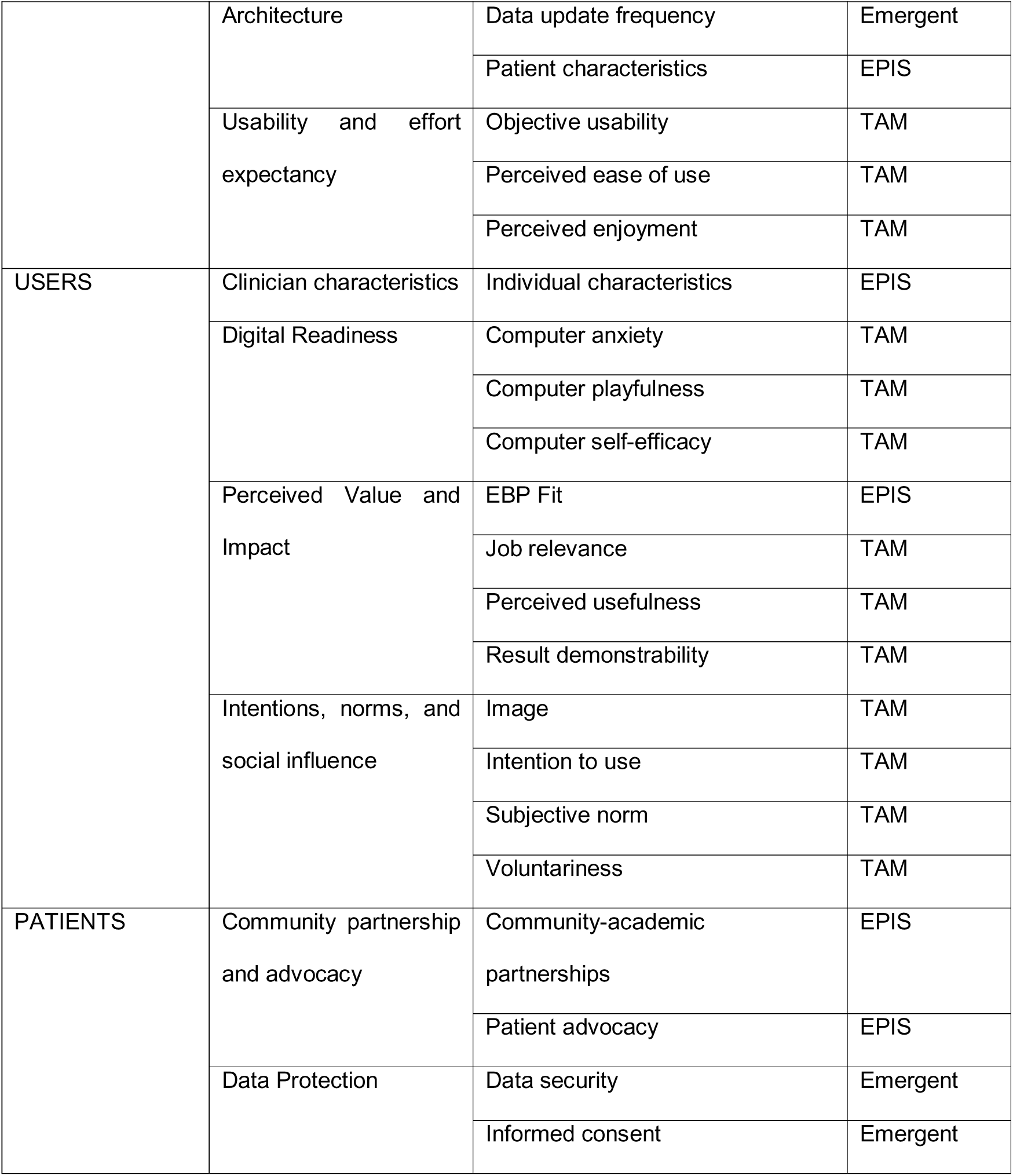
The 36 codes, their thematic domain, and theoretical provenance (EPIS, TAM, or emergent).

Within this integrated coding framework, Perceived Usefulness and Organizational Characteristics emerged as the most prevalent and centrally connected codes across stakeholder groups, reflecting their cross-cutting relevance. Network visualizations (Figure 1) illustrate the relative prominence and co-occurrence of codes across stakeholder groups. Panel I (Decision-makers) highlights Governance and Organizational Characteristics as dominant nodes; Panel II (Experts) shows dense clustering around Organizational Characteristics and Experience; in Panel III (Users), Perceived Usefulness emerges as the most salient code, closely linked to organizational constraints. Finally, Panel IV (Patient organizations) is characterized by the prominence of Patients’ Advocacy.

**Figure 1.**
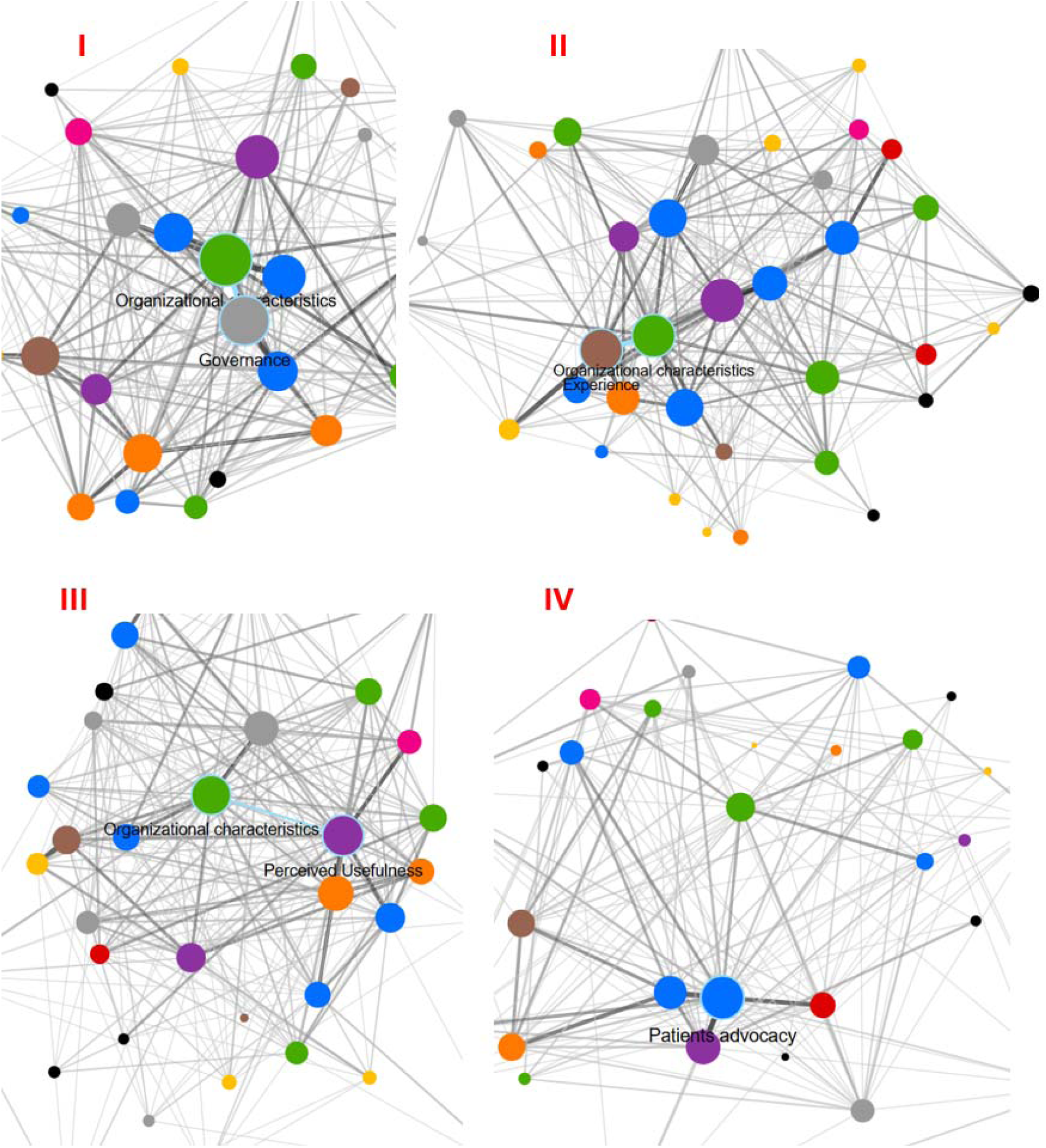
Network visualization of code co-occurrence across stakeholder groups. Network graphs depict the relative prominence and co-occurrence of qualitative codes derived from thematic analysis of 29 interviews and focus groups. Node size reflects code frequency, and edge density represents the strength of co-occurrence between codes. Panel I corresponds to decision-makers; Panel II to clinical and technical experts; Panel III to end users (hospital-based clinicians); and Panel IV to patient organizations. Across the full dataset, Perceived Usefulness and Organizational Characteristics emerged as the most central and interconnected codes. At the same time, group-specific patterns highlight the emphasis on Governance among decision-makers, Experience among experts, Perceived Usefulness among users, and Patients’ Advocacy among patient organizations.

Following this group-level analysis, the 15 categories were further synthesized into four overarching themes that structure the findings: Environment, Technical Dimension, Users, and Patients. Figure 2 (Sankey diagram) illustrates the relative distribution of these 15 categories across stakeholder groups, highlighting both shared determinants and group-specific emphases. Representative quotations organized by theme are presented in Table 2. Quotations are labeled by stakeholder group and data collection format: 100-series (decision-makers), 200-series (experts), 300-series (users), and 400-series (patient organizations); the number after the hyphen denotes interview (1) or focus group (2).

**Figure 2.**
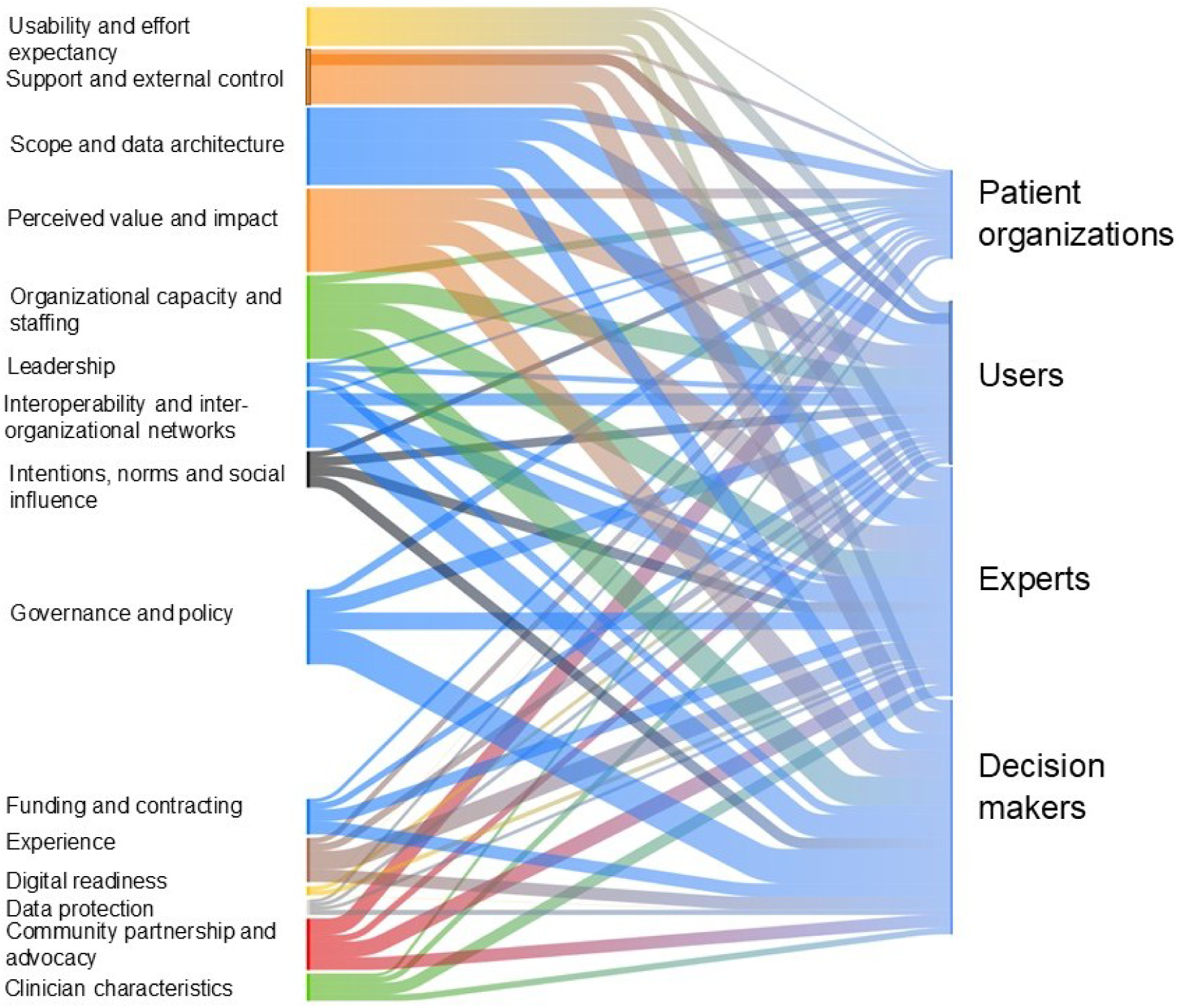
Sankey diagram showing the integration of analytic categories across stakeholder groups. The diagram illustrates how 15 higher-order categories, derived from 36 qualitative codes, are distributed across stakeholder groups. Flow width represents the relative salience of each category within a group. The visualization highlights both shared and group-specific emphases, demonstrating how environmental, technical, user-level, and patient-centered determinants intersect to shape implementation of a national IEI registry.

**Table 2.**
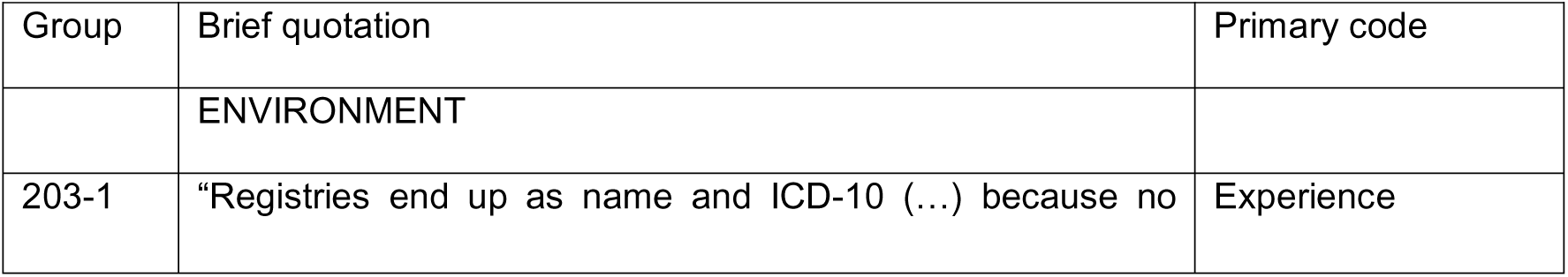
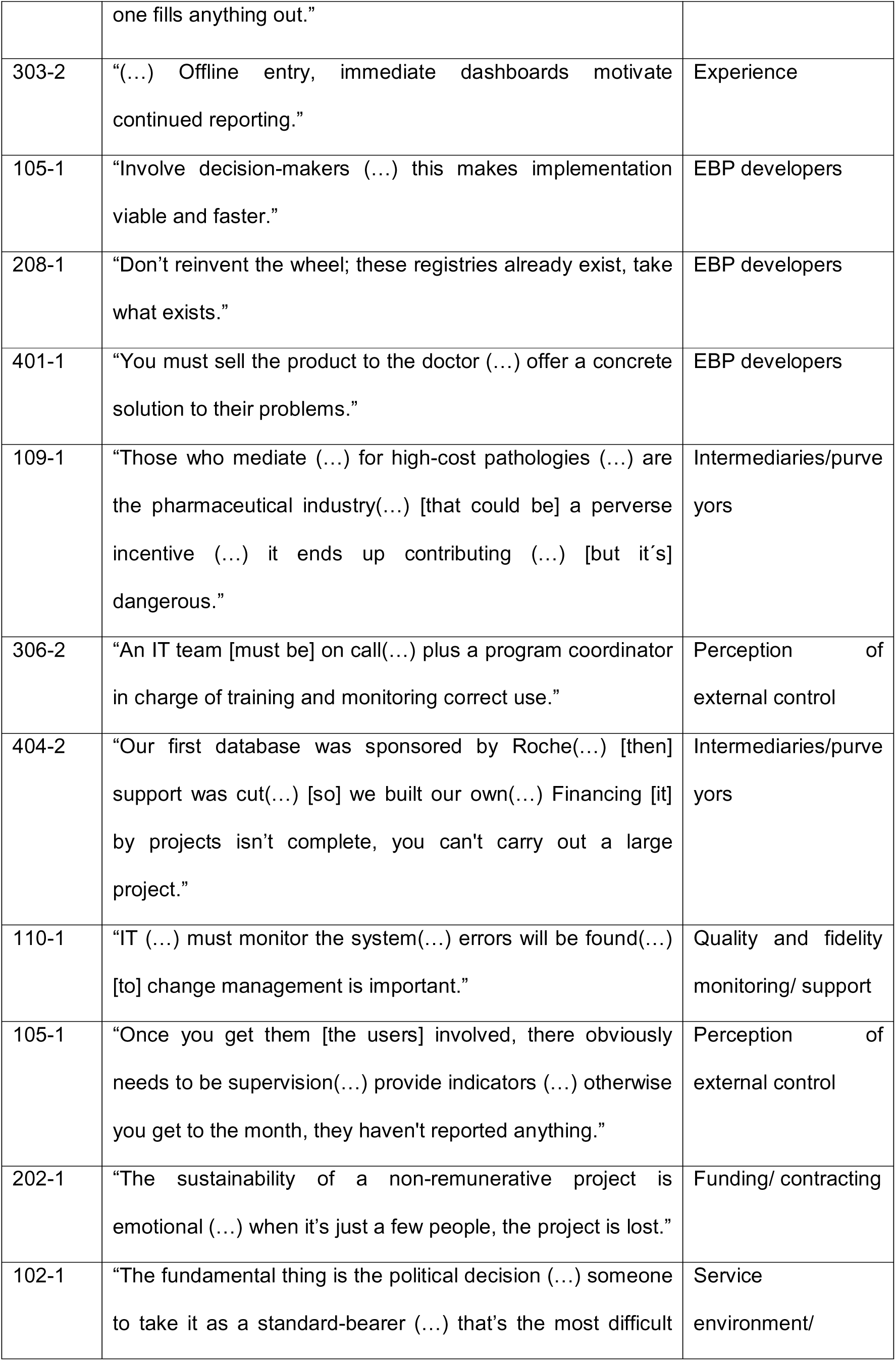
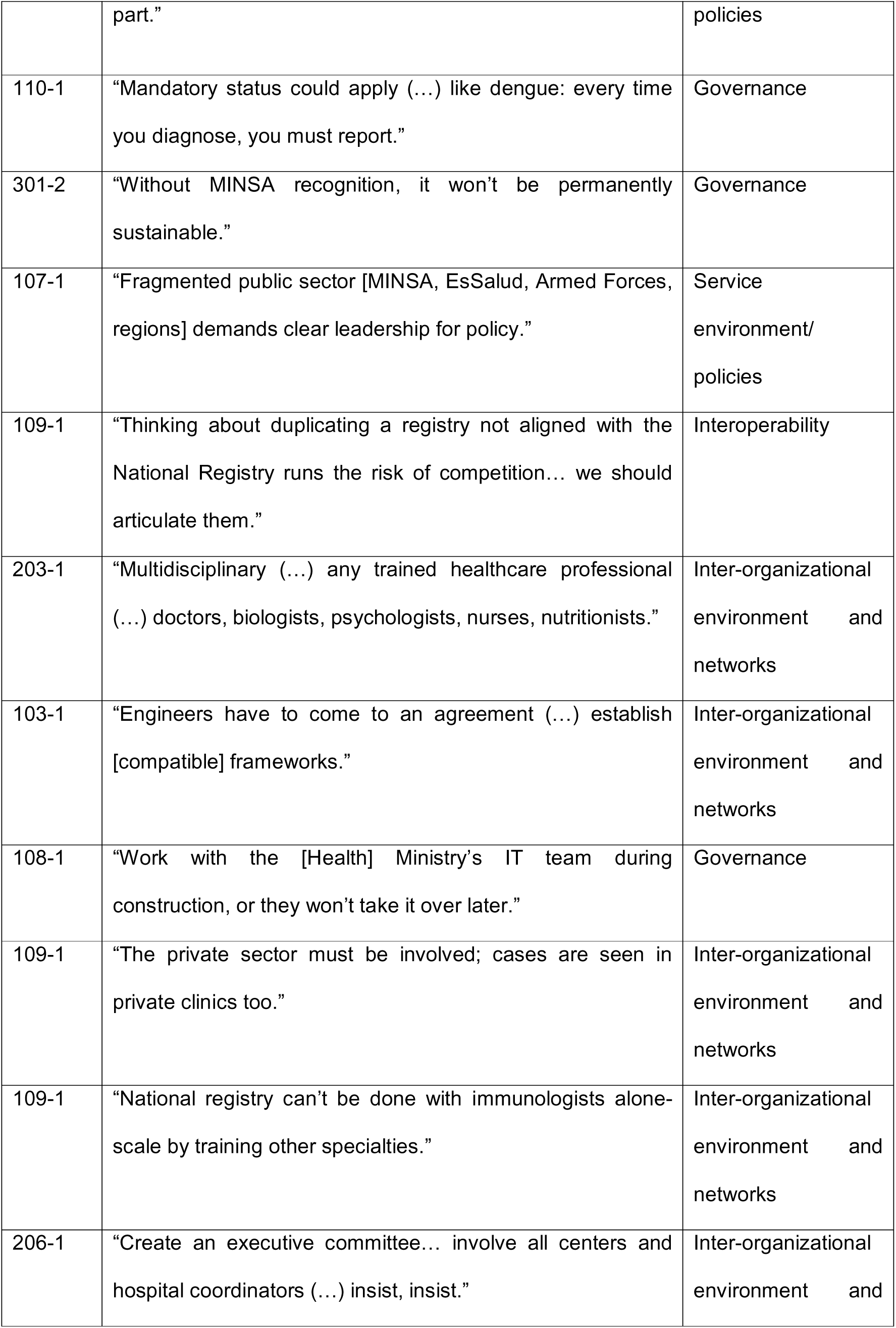
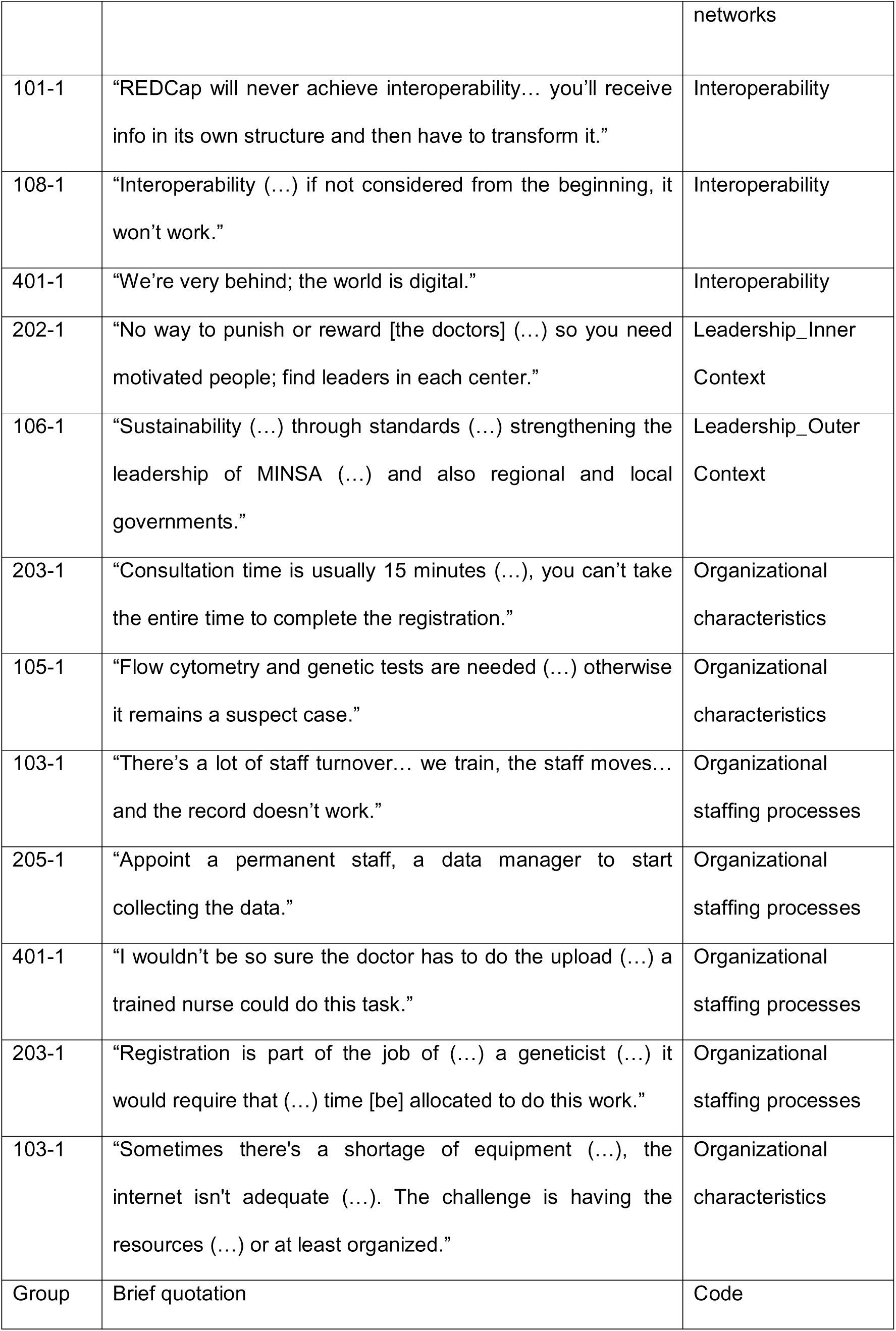
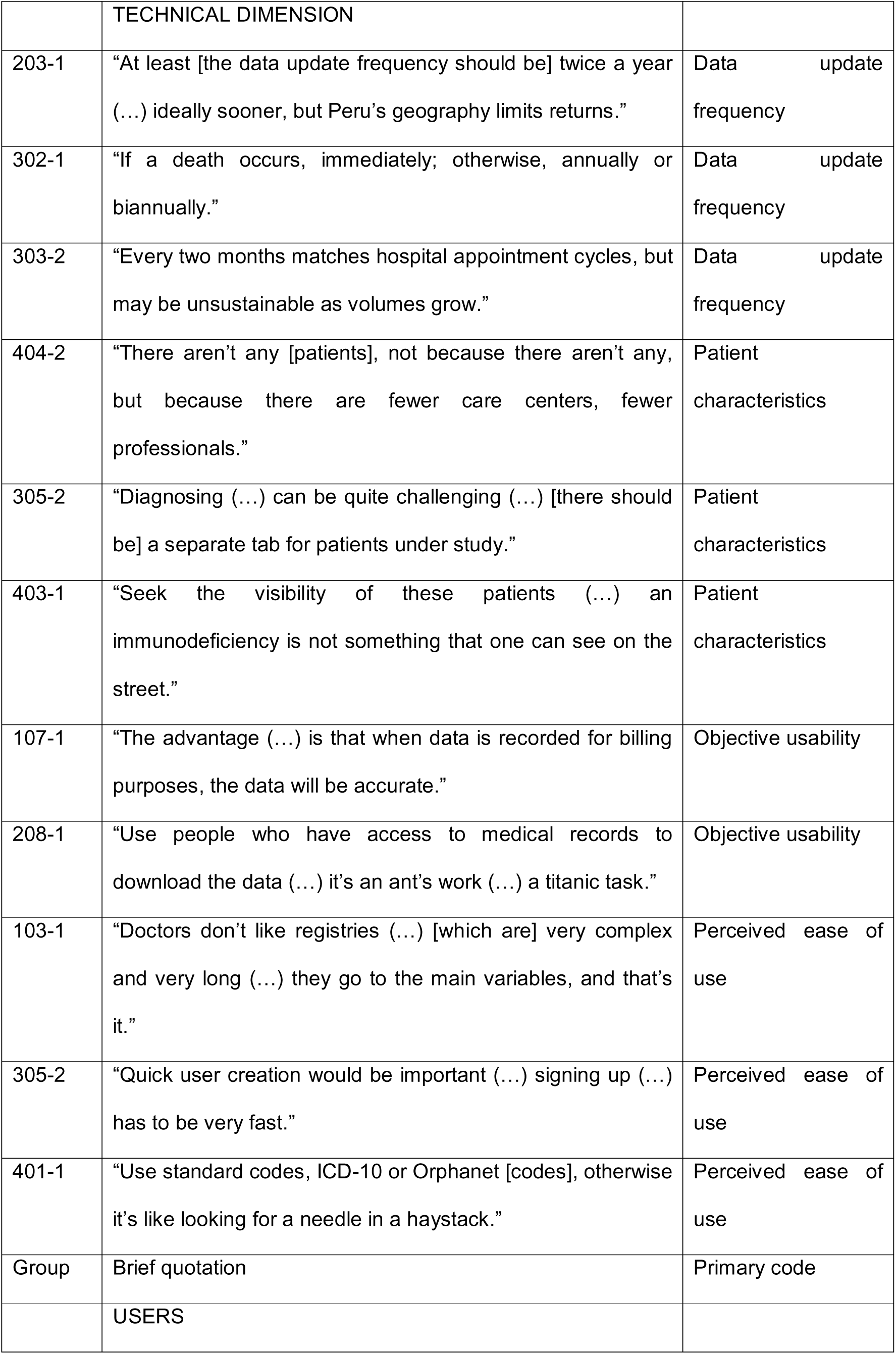
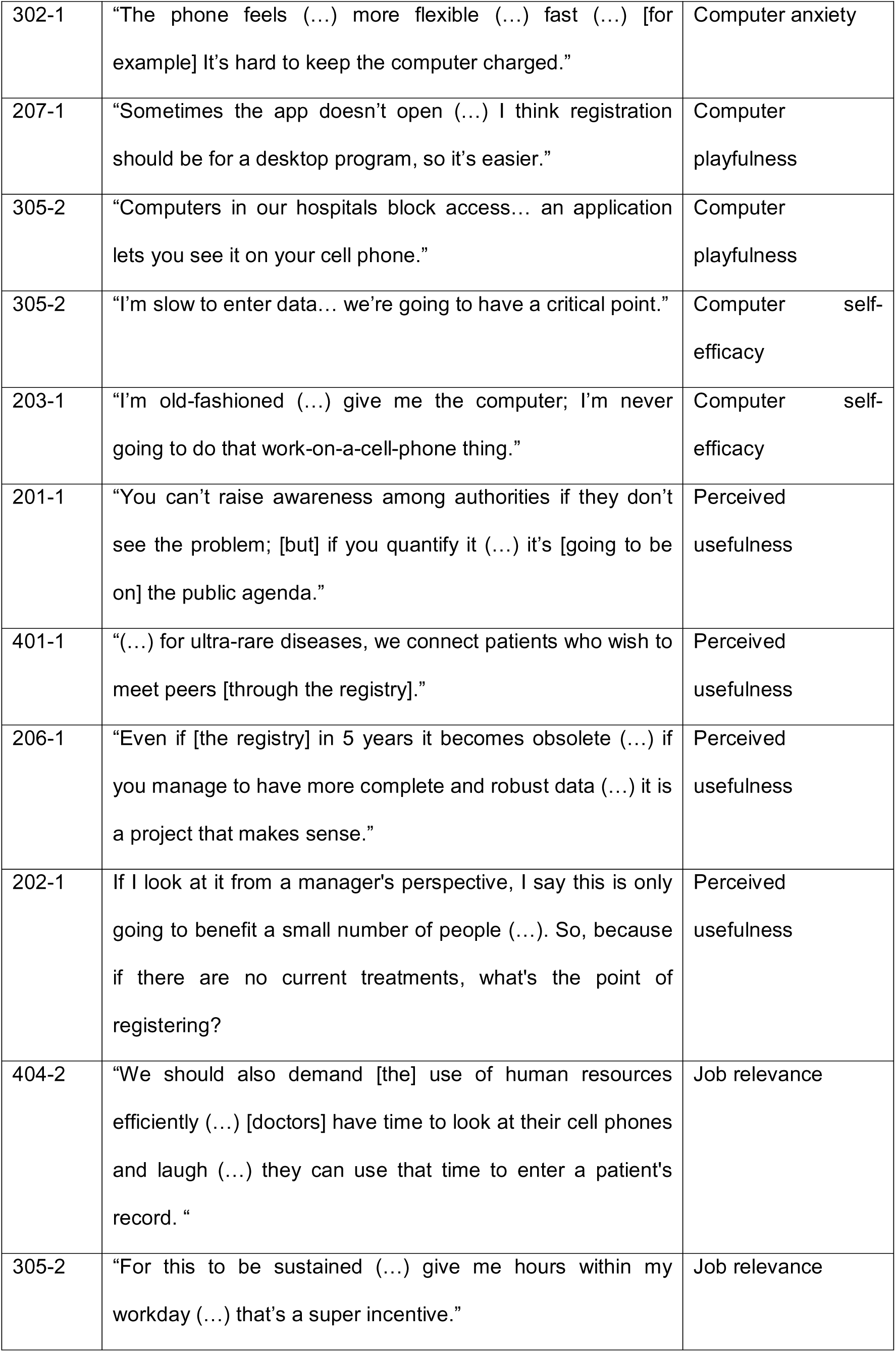
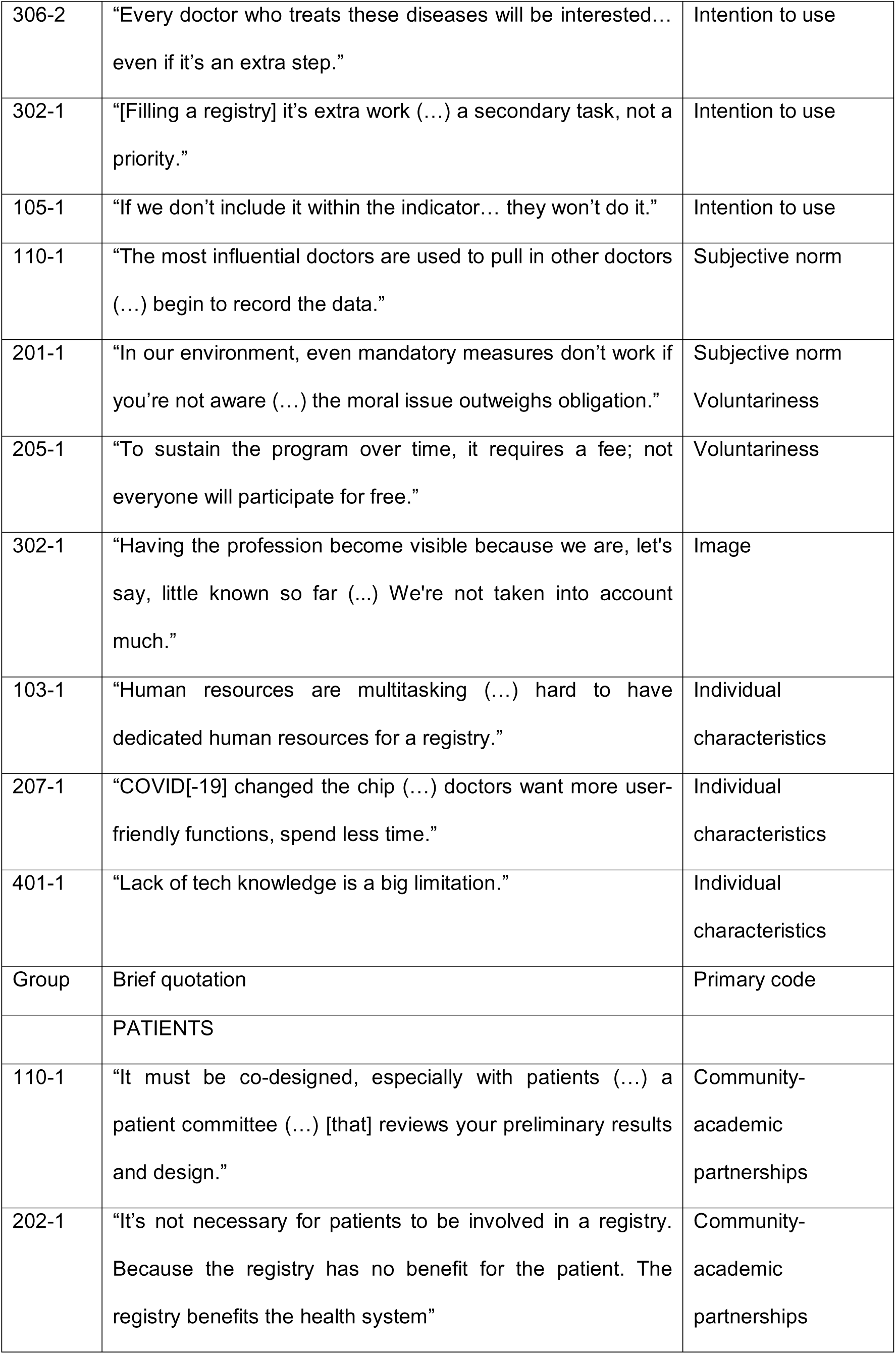
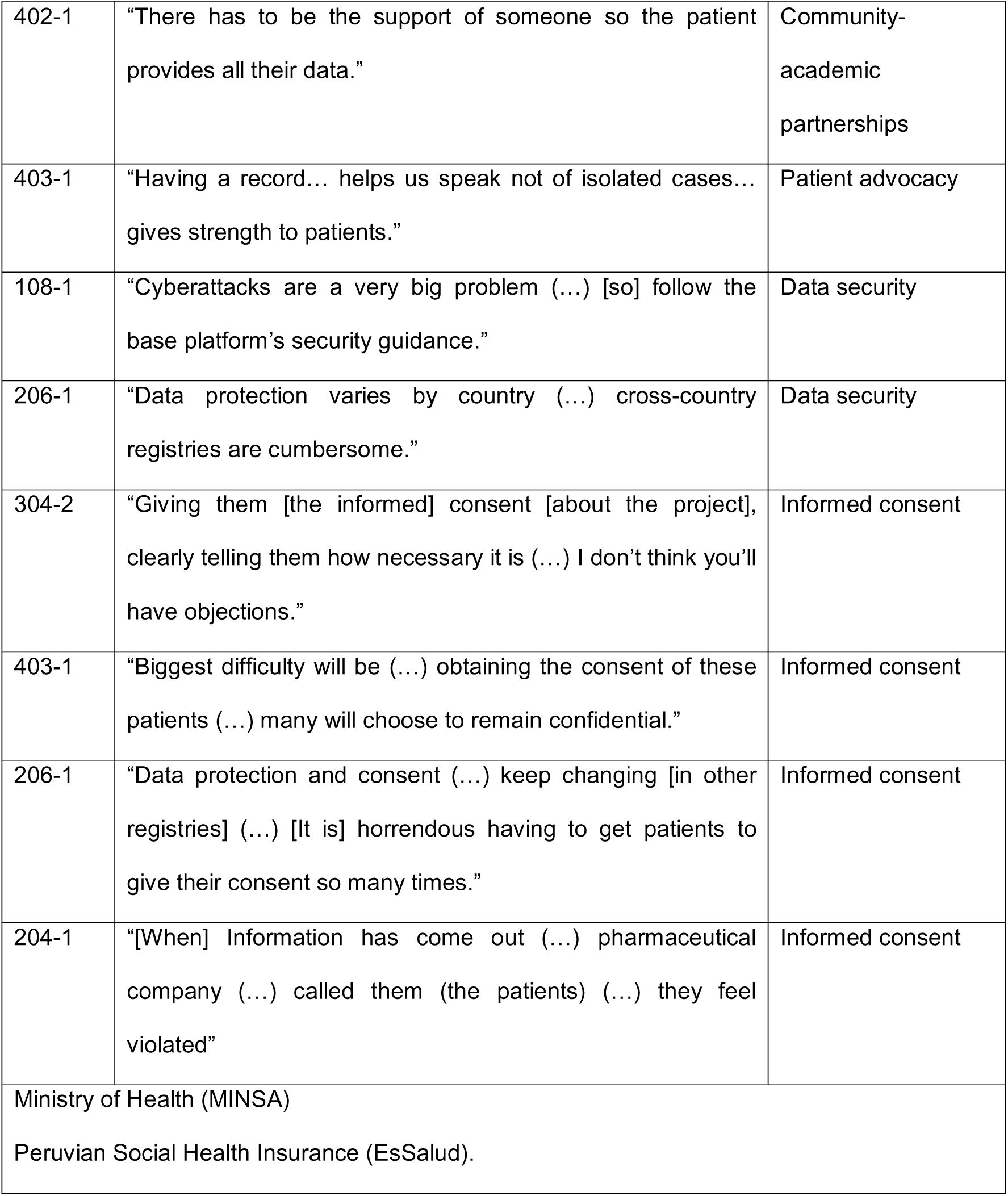
Selected illustrative quotations by theme.

### Environment

Governance was frequently mentioned, including legal authority, ownership, leadership structures, and continuity amid staff turnover. Participants emphasized the importance of formal recognition and policy backing. One decision-maker noted that *“the fundamental thing is the political decision (…) someone to take it as a standard-bearer”* (102-1). Regulatory anchoring was also linked to reporting expectations; as one decision-maker explained, *“Mandatory status could apply (…) like dengue: every time you diagnose, you must report”* (110-1). Stakeholders further described the need for registry activities to be incorporated into formal roles rather than relying on voluntary effort, with one expert warning that *“the sustainability of a non-remunerative project is emotional (…) when it’s just a few people, the project is lost”* (202-1).

Participants also referred to challenges related to health system fragmentation and variability in institutional capacity. Organizational constraints were also highlighted. One expert noted that *“consultation time is usually 15 minutes (…), you can’t take the entire time to complete the registration”* (203-1), while another decision-maker reported that *“there’s a lot of staff turnover… we train, the staff moves… and the record doesn’t work”* (103-1). Resource limitations, including shortages of equipment, internet access, and diagnostic capacity, were repeatedly mentioned as part of the implementation context.

Interoperability and organizational capacity were also described as recurrent implementation conditions. Participants referred to the use of standardized coding systems, patient identifiers, and alignment with existing systems, and they stressed that interoperability should be considered from the outset. One decision-maker stated that *“Interoperability (…) if not considered from the beginning, it won’t work”* (108-1), while another warned that duplicating a registry *“not aligned with the National Registry runs the risk of competition”* (109-1). Coordination across institutions and disciplines was also emphasized. One expert suggested to *“create an executive committee… involve all centers and hospital coordinators”* (206-1), and another expert described the registry as requiring a multidisciplinary network involving *“doctors, biologists, psychologists, nurses, [and] nutritionists”* (203-1). Participants additionally referred to the need for technical support and designated oversight roles, including IT monitoring, training, and supervision.

### Technical Dimension

Participants described technical requirements related to data structure, usability, and system functionality. A recurring topic was the definition of a minimal dataset including patient identification, diagnostic categories, clinical manifestations, laboratory and genetic data, treatments, and follow-up information. At the same time, participants warned against overly complex designs. One decision-maker noted that *“when data is recorded for billing purposes, the data will be accurate”* (107-1), while one expert emphasized the workload involved in retrospective abstraction, describing it as *“an ant’s work (…) a titanic task”* (208-1). Concerns were also raised about the scope of variables and the need to accommodate diagnostic uncertainty, with one user suggesting *“a separate tab for patients under study”* (305-2). Participants also referred to update frequency, proposing periodic updates combined with event-based reporting; for example, one expert suggested that updates should occur *“at least twice a year (…) ideally sooner, but Peru’s geography limits returns”* (203-1), while one user noted that *“every two months matches hospital appointment cycles, but may be unsustainable as volumes grow”* (303-2).

Usability-related aspects included system speed, interface simplicity, and integration with existing workflows. Participants consistently described preferences for short data entry times and interfaces that support routine clinical use. One decision-maker stated that *“Doctors don’t like registries (…) [which are] very complex and very long”* (103-1), and one user added that *“signing up (…) has to be very fast”* (305-2). Standardized coding was also identified as a practical usability issue; as one patient organization representative put it, *“Use standard codes, ICD-10 or ORPHANET, otherwise it’s like looking for a needle in a haystack”* (401-1). Mobile access was mentioned for quick consultation in other parts of the corpus, whereas desktop-based entry was more often associated with full documentation tasks. Challenges such as data duplication and limited interoperability across systems were also reported.

Quantitative prioritization of variables was conducted using Likert-scale ratings from experts and users, and results were integrated with qualitative findings (Supplementary Table S3). Additionally, differing perspectives on specific technical design elements were documented, including the scope of variables, handling of suspected versus confirmed cases, and user access roles (Supplementary Table S4).

### Users

Participants described factors influencing adoption at the user level, including time availability, digital skills, and perceived usefulness. Time constraints were frequently reported, particularly regarding clinical workload and documentation burden. One user described registry completion as *“extra work (…) a secondary task, not a priority”* (302-1), while another emphasized that sustainability would require *“hours within my workday”* (305-2). However, this view was not uniform across groups. A patient organization representative argued that registry use also depends on how existing time is organized, stating that *“[doctors] have time to look at their cell phones and laugh (…) they can use that time to enter a patient’s record”* (404-2). Stakeholders also discussed the feasibility of shorter forms and distributed workflows involving multiple roles.

Digital familiarity varied among participants, with some indicating preferences for specific interface types depending on the task. One expert stated, *“I’m old-fashioned (…) give me the computer”* (203-1), whereas one user noted that *“computers in our hospitals block access… an application lets you see it on your cell phone”* (305-2). Perceived usefulness was discussed in relation to outputs such as reports, dashboards, or institutional benefits; one decision-maker noted that if the problem is quantified, *“it’s [going to be on] the public agenda”* (201-1), while one expert stated that *“more complete and robust data (…) [make] it a project that makes sense”* (206-1). Participants also referred to the influence of peers and institutional practices, with one decision-maker noting that *“The most influential doctors are used to pull in other doctors”* (110-1). Different views were expressed regarding voluntary versus mandatory participation, as well as the role of institutional support and protected time.

### Patients

Participants described the roles of patients and patient organizations in registry development and implementation. References were made to their participation in advisory or governance structures, as well as their role in advocacy and awareness. One decision-maker stated that the registry *“must be co-designed, especially with patients (…) a patient committee (…) [that] reviews your preliminary results and design”* (110-1). At the same time, views were not uniform across groups: one expert argued that *“it’s not necessary for patients to be involved in a registry”* because *“the registry benefits the health system”* (202-1), whereas a patient organization representative emphasized that *“having a record… helps us speak not of isolated cases… gives strength to patients”* (403-1). Stakeholders thus discussed the balance between inclusion of patient perspectives and the definition of clinically relevant data.

Data protection and informed consent were consistently mentioned. Participants referred to mechanisms such as anonymization, role-based access, and institutional oversight. One decision-maker warned that *“Cyberattacks are a very big problem”* (108-1), while one expert noted that *“data protection varies by country (…) cross-country registries are cumbersome”* (206-1). Communication about data use and safeguards was also described as part of the process; one user suggested that by *“clearly telling them how necessary it is”* patients would be more willing to participate (304-2). At the same time, patient organizations highlighted confidentiality concerns, with one stating that the *“biggest difficulty”* would be obtaining consent because *“many will choose to remain confidential”* (403-1), and one expert recalling that repeated consent requirements in other registries had become *“horrendous”* (206-1). Participants also referred to the use of registry data to support visibility of rare conditions and inform decision-making processes.

Finally, Figure 3 presents a simplified conceptual model linking the four overarching themes (Environment, Technical Dimension, Users, and Patients). The original detailed version of this pathway is provided in Supplementary Figure S1. The figure maps the categories within these four themes and shows the relationships identified between them.

**Figure 3.**
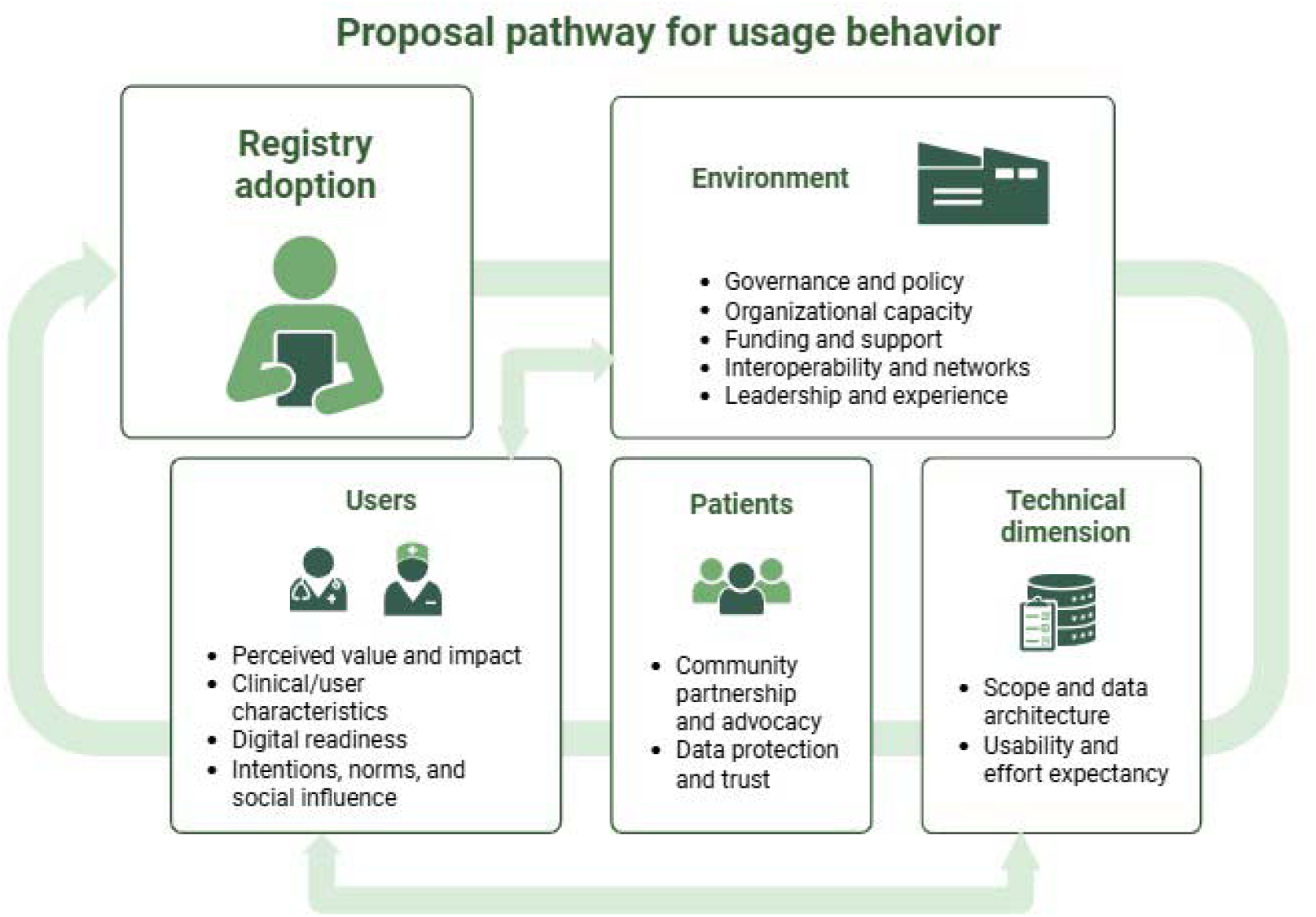
Simplified pathway for registry adoption integrating EPIS- and TAM-informed dimensions. The figure organizes the findings into four interrelated dimensions; Environment, Users, Patients, and Technical Dimension, linked to registry adoption. The arrows indicate the dynamic and interdependent relationships among these dimensions in shaping registry adoption and use. The original detailed version of this conceptual pathway is provided in Supplementary Figure S1. Created with <u>BioRender.com</u>

## Discussion

This Phase I study aimed to identify the requirements, priorities, and implementation conditions for the development of a national registry for inborn errors of immunity (IEI) in Peru from the perspective of key stakeholders. It identified four interrelated dimensions shaping the prospective implementation of a national registry for IEI in Peru: Environment, Technical Dimension, Users, and Patients. Together, these findings indicate that registry adoption is not determined by the digital platform alone, but by the alignment between governance arrangements, sociotechnical design, workforce realities, and patient trust.

In our coding structure, Perceived Usefulness and Organizational Characteristics emerged as highly central constructs across stakeholder groups, while the thematic synthesis showed that implementation determinants operate simultaneously at system, organizational, individual, and patient levels. This pattern is consistent with the logic of implementation science frameworks, which emphasize that uptake and sustainability depend on the interaction between the intervention and its context rather than on technical adequacy alone. The environmental dimension highlights the central role of governance, policy backing, financing, and inter-organizational coordination. Participants repeatedly described the registry as unlikely to be sustainable without formal recognition by the Ministry of Health, defined ownership, and explicit assignment of responsibilities for reporting, maintenance, and oversight.

In a health system as segmented as Peru’s, where care is distributed across MINSA, EsSalud, the armed forces, police services, and the private sector, these findings reinforce the idea that registry implementation is primarily a governance-dependent process. This aligns with evidence from health systems and implementation research showing that digital health initiatives in low- and middle-income countries (LMICs) frequently fail or stagnate when they are not embedded in stable institutional arrangements or when they depend on isolated champions, short-term funding, or poorly coordinated actors (9,14,18,31). Our results, therefore, suggest that a national IEI registry should be framed not merely as a data repository, but as a health system function requiring policy endorsement, organizational ownership, and mechanisms for continuity across institutional boundaries. This also supports the need to articulate the registry with broader national efforts on rare and orphan diseases, so that it can contribute not only to clinical follow-up and service coordination, but also to visibility, monitoring, and evidence-informed decision-making for these conditions. Ultimately, sustaining a truly national IEI registry will depend on the political decision to recognize and support it as a strategic health system function.

The technical dimension revealed a consistent tension between data comprehensiveness and operational feasibility. Stakeholders favored a structured but limited minimum dataset that could support meaningful clinical follow-up while remaining feasible under real-world workload conditions. This reflects broader rare disease registry guidance, which recommends parsimonious common data elements, interoperability-oriented architecture, and staged data collection to improve feasibility and data quality (26,32,33). Participants also emphasized that interoperability was not only a desirable technical feature but a necessary mechanism to reduce duplication, lessen documentation burden, and improve data quality in a health system characterized by fragmented clinical and information flows. This interpretation is aligned with digital health literature showing that interoperability is foundational for effective digital medicine (34), and with work underscoring the importance of standardized classifications and ontologies for data harmonization and precision-oriented health information systems (35). In rare disease settings, interoperable approaches have also been proposed as a way to enable distributed data quality assessment across fragmented hospital information systems (36).

Usability was similarly described as inseparable from implementation success. Interface simplicity, speed, reduced duplication, and compatibility with existing workflows were repeatedly described as necessary conditions for routine use. These observations are consistent with sociotechnical research showing that digital tools are more likely to be adopted when they reduce friction within work processes rather than adding parallel layers of documentation (17). In this sense, the technical findings of Phase I do not simply specify what the registry should contain; they also define the boundaries of what can be realistically, operationally feasible, and sustainable in the Peruvian context. The selection of REDCap for the prototype is also coherent with its established use as a metadata-driven platform for research data capture and workflow support (37). The challenge is not unique to Peru. Even well-established IEI and rare disease registries have required progressive refinement of case definitions, common data elements, governance arrangements, and interoperability strategies to remain usable across institutions and over time (10,12,13,26,33). For Peru, where the health system is fragmented and digital maturity is uneven, these tensions are likely to be even more pronounced and reinforce the need for a context-adapted, phased implementation strategy.

The user dimension strongly supports the relevance of a TAM-informed interpretation. Participants repeatedly linked intended use to the anticipated value of the registry, especially whether it would generate outputs such as dashboards, clinical follow-up information, institutional visibility, research opportunities, or policy leverage. This is consistent with prior Peruvian evidence from the National Death Information System (SINADEF), where physicians’ perceived usefulness and perceived ease of use predicted future intention to use a national digital health information system (24). More broadly, these findings are aligned with the health informatics literature showing that technology adoption by clinicians is shaped not only by perceived usefulness and ease of use, but also by workflow fit, organizational context, and implementation conditions(29,38).

In our study, perceived usefulness was clearly embedded in organizational realities. Users did not evaluate the registry in abstraction; they evaluated it against competing clinical demands, protected time, staffing constraints, access to devices, institutional expectations, and the practical contribution the registry could make to clinical follow-up and research. Social and organizational cues were also mentioned, particularly influential peers, performance indicators, and the need for leadership endorsement. These findings suggest that TAM constructs in this setting are not purely individual attitudes but are mediated by organizational conditions and work design (17,29,38). They also suggest that adoption is closely tied to concrete design choices, including a minimum data set that is feasible in practice, a user-centered interface, and workflow strategies that reduce rather than increase documentation burden. Thus, adoption depends on whether users can see both the value and the practical possibility of using the registry within routine care.

The patient dimension adds an essential layer of legitimacy, trust, and social value. Participants described patient organizations as important for advocacy, visibility, and awareness, while also expressing caution about the extent and form of patient involvement in registry governance and design. The contrasting views observed across stakeholder groups suggest that this tension is a recurring feature of rare disease registries, where public value, stakeholder participation, and feasible governance must be negotiated in practice (26,39). Data protection and informed consent emerged as particularly salient, with concerns related to confidentiality, institutional trust, and the ethical management of identifiable or sensitive information. These observations are consistent with rare disease registry guidance emphasizing that public value depends not only on data completeness, but also on robust privacy protections, transparent governance, and communication that sustains patient confidence (12,26). In this study, trust was not a secondary ethical issue; it was an implementation condition.

The combined use of EPIS and TAM was deliberate and methodologically coherent with the aims of Phase I. Among implementation frameworks, EPIS was particularly suitable because this project was explicitly organized as a staged implementation process and required simultaneous attention to outer-context conditions, such as governance, policy, financing, and inter-organizational relationships, and inner-context determinants, such as leadership, staffing, workflow integration, and organizational readiness (18). TAM, in turn, was selected because a core analytic concern of this phase was not only implementation feasibility but also intention to use among clinical users(29,38). TAM offered a parsimonious and highly interpretable lens centered on perceived usefulness, perceived ease of use, and behavioral intention, which aligned closely with both the interview data and prior Peruvian evidence from SINADEF (24). In this study, EPIS helped explain “where” implementation determinants were located, whereas TAM helped explain “how” users interpreted the prospective value and effort of the registry. Figure 3 summarizes a multilevel implementation pathway in which environmental governance conditions shape technical design, technical design conditions user acceptance, and patient-facing legitimacy influences the social sustainability of registry use. This is consistent with the broader literature on digital health governance in LMICs (14,31).

This study has several strengths. It incorporated perspectives from four stakeholder groups spanning policy, clinical expertise, frontline use, and patient advocacy, allowing triangulation across multiple levels of the health system. It also combined qualitative inquiry with structured prioritization of data elements, strengthening the translation of stakeholder perspectives into early design outputs. The use of EPIS and TAM further provided complementary interpretive lenses that connected governance and implementation context with user-level acceptance and intention to use. An additional strength is that the study was conducted in Peru, where health system fragmentation and uneven institutional capacity shape real-world feasibility, whereas most established IEI and rare disease registries have been developed and reported in higher-income settings. The identification of shared themes across different health subsystems is also a relevant strength, as it reveals areas of convergence that may facilitate future coordination through the registry.

However, several limitations should also be acknowledged. First, although the sample intentionally captured diverse stakeholder groups, the findings are grounded in the Peruvian context and may not fully represent all regional realities beyond the institutions and networks included in this phase. Second, the study explored anticipated implementation rather than observed behavior; therefore, conclusions regarding adoption and sustained use remain provisional and should be tested in real usability and implementation settings. Third, because the registry was still in the design phase, some views were shaped not only by expectations about the proposed IEI registry itself but also by participants’ prior experiences with other digital health systems, such as electronic health records or registries used for other conditions. This does not alter the empirical findings of this phase, but it does qualify their interpretation, as some anticipated benefits, concerns, and critiques may reflect transferred experience rather than direct interaction with a fully functional IEI registry. Fourth, while patient organizations were included, the perspectives of individual patients or caregivers outside organized advocacy structures were less represented. Finally, because this phase intentionally integrated qualitative findings with structured variable prioritization before direct usability testing, some technical preferences and design priorities may evolve once users interact directly with the prototype under realistic task conditions.

Taken together, these findings strengthen the argument that the implementation of an IEI registry in Peru should be approached as a context-sensitive digital health intervention rather than as a simple adaptation of an existing registry model or a mere data repository. The main contribution of Phase I lies not only in identifying which data elements stakeholders consider relevant, but also in defining the governance, usability, organizational, and ethical conditions under which those data can be collected and sustained. This provides a stronger foundation for Phase II usability testing and, subsequently, for feasibility assessment under real-world health system conditions. More broadly, it also aligns with calls in rare disease research for interoperable, well-governed, and reusable data infrastructures that can expand the value of scarce and distributed clinical information (39).

## Data Availability

All data produced are available online at https://zenodo.org/records/20614353

## SUPPLEMENTARY INFORMATION

S1 Table. COREQ checklist.

S2 Table. Description of participants.

S3 Table. Mapping of EBP characteristics through mixed-methods triangulation.

S4 Table. Divergent stakeholder perspectives on key technical design decisions.

**S1 Figure.**
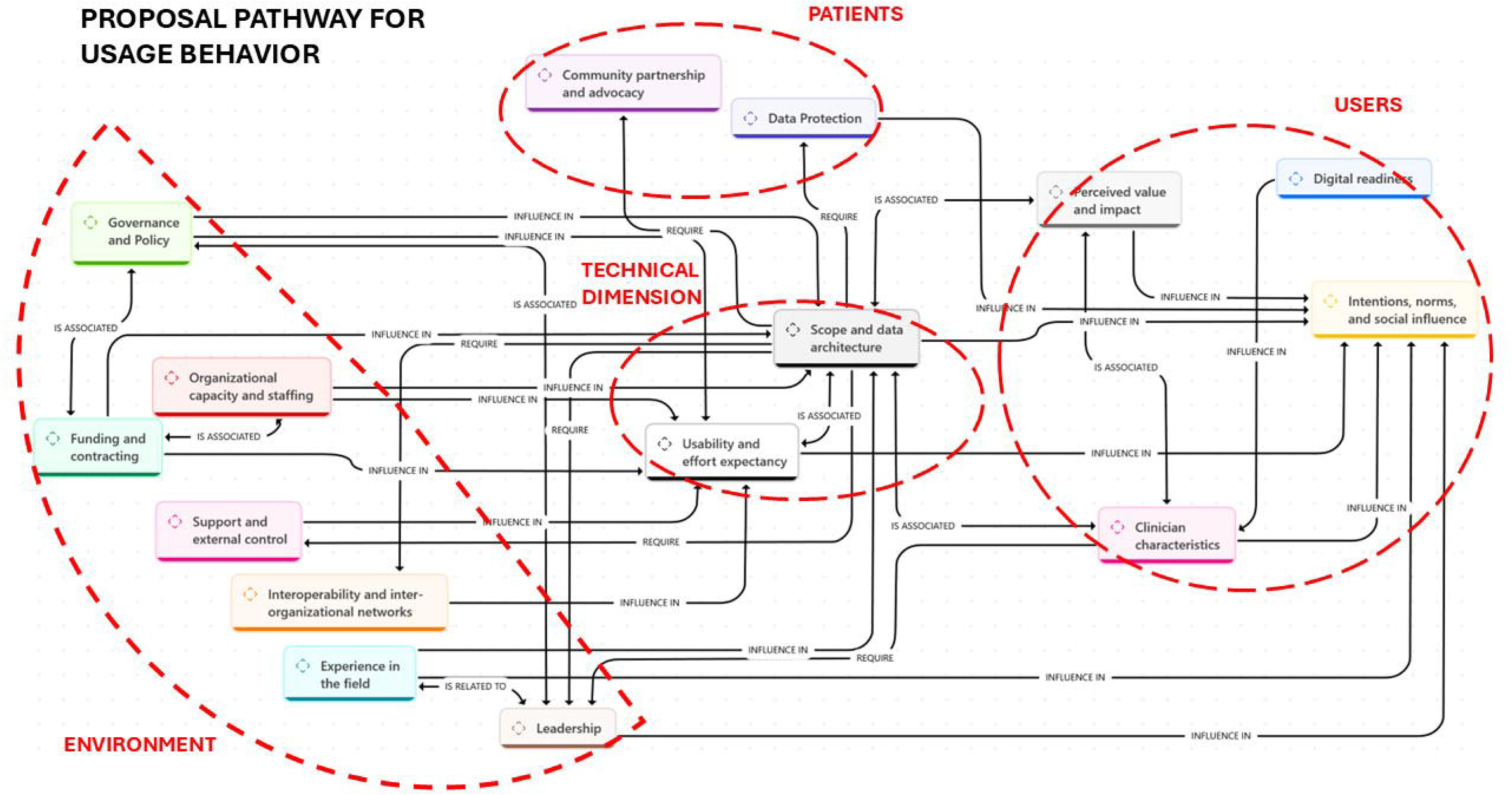
Original detailed conceptual pathway linking the four thematic dimensions and category-level relationships.

