## Supplementary tables and figure for "Toward a National Registry for Inborn Errors of Immunity in Peru: A Qualitative Implementation Study"

**Supplementary Table S1.** Consolidated Criteria for Reporting Qualitative Research (COREQ) checklist

| Item | COREQ criterion | Description in this study |
| --- | --- | --- |
| Domain 1: Research team and reflexivity | | |
| Personal Characteristics | | |
| 1 | Interviewer/facilitator | All interviews and focus groups were conducted by the principal investigator and a psychologist. |
| 2 | Credentials | 1. Principal investigator: MD, PhD candidate in Health Sciences.  2. Psychologist trained in qualitative methods. |
| 3 | Occupation | 1. Physician–researcher affiliated with a public university and national research projects.  2. Psychologist–researcher affiliated with a public university and national research projects. |
| 4 | Gender | Both females. |
| 5 | Experience and training | 1. Formal training in clinical immunology and clinical research, prior experience in inborn error of immunity.  2. Formal training in qualitative methods, prior experience conducting interviews and focus groups. |
| Relationship with participants | | |
| 6 | Relationship established | 1. Prior professional relationship with group III (Users).  2. No prior personal relationship with most participants; some professional awareness existed for experts and decision-makers. |
| 7 | Participant knowledge of interviewer | Participants were informed of the interviewer’s academic role, research objectives, and non-regulatory position. |
| 8 | Interviewer characteristics | 1. Clinical background in immunology and involvement in registry development acknowledged  2. Research background in qualitative studies, analytic rigor strengthened through triangulation and transparent coding. |
| Domain 2: Study design | | |
| Theoretical framework | | |
| 9 | Methodological orientation | Phenomenology with hybrid thematic analysis guided by EPIS and TAM, with inductive coding for emergent constructs. |
| Participant selection | | |
| 10 | Sampling | Purposive sampling to capture heterogeneity across policy, clinical, technical, and patient perspectives. |
| 11 | Method of approach | Participants were contacted via telephone, email and institutional coordination. |
| 12 | Sample size | 29 qualitative sessions (in-depth interviews and focus groups). |
| 13 | Non-participation | Non-participation was due to any response to the team’s messages, and scheduling constraints; in total were 18 refusals. |
| Setting | | |
| 14 | Setting of data collection | Virtual (videoconference), or in-person (coffee shops, office, and hospital settings). |
| 15 | Presence of non-participants | No non-participants were present during data collection. |
| 16 | Description of sample | Decision-makers, experts, hospital-based users, and patient organizations (Supplementary Table S2). |
| Data collection | | |
| 17 | Interview guide | Semi-structured guides based on predefined domains; developed and reviewed by six content experts |
| 18 | Repeat interviews | No repeat interviews were conducted. |
| 19 | Audio/visual recording | Sessions were audio-recorded with participant consent when they were in person, and video-recorded with participant consent when they were virtual. |
| 20 | Field notes | Reflexive and contextual field notes were taken during and after sessions. |
| 21 | Duration | Sessions typically lasted approximately 45 minutes, with a small number extending to 90 minutes. |
| 22 | Data saturation | Thematic saturation was achieved across stakeholder groups by the 28th session. However, one additional interview with a decision-maker had already been scheduled and was completed, generating relevant insights that were incorporated into the analysis. |
| 23 | Transcripts returned | Transcripts were not returned to participants for checking. |
| Domain 3: Analysis and findings | | |
| Data analysis | | |
| 24 | Number of data coders | Primary coding was conducted independently by the principal investigator and a psychologist, both of whom participated in data collection. Interview transcripts were translated into English and reviewed by both coders and by a senior supervisor from the University of Cincinnati, who served as an external auditor. This supervisor also provided oversight on framework alignment, coding consistency, and analytic rigor, ensuring quality and reliability throughout the analytic process. |
| 25 | Coding tree | The coding tree comprised 36 codes organized into 15 categories and four overarching themes, based on the final, iteratively refined version of the codebook (v4.3) (Table 1). |
| 26 | Derivation of themes | Themes were derived through a hybrid deductive–inductive approach, combining framework-informed coding with inductive theme generation. |
| 27 | Software | ATLAS.ti was used for data management and coding. |
| 28 | Participant checking | Formal member checking of transcripts or themes was not performed. However, preliminary findings were presented to user participants through in-person and virtual sessions as part of preparation for Phase II, which will be conducted with their active participation. Credibility was further strengthened through methodological triangulation. Progressive dissemination of findings to other stakeholder groups is planned. |
| Reporting | | |
| 29 | Quotations presented | Representative quotations presented and identified by stakeholder group and session code. |
| 30 | Data and findings consistency | Findings were consistent with supporting quotations. |
| 31 | Clarity of major themes | Four overarching themes clearly presented. |
| 32 | Clarity of minor themes | Codes and categories were explicitly reported (e.g., Table 3). After finalization of the codebook (v4.3), peer coding was conducted under the supervision of an external auditing researcher. Once coding was completed, categories were established through team-based discussion and reviewed with senior researchers experienced in qualitative research. Divergent perspectives were resolved through analytic consensus. The final themes were subsequently refined and validated through the same review process, including examination of relationships across themes that informed the final integrative model presented in the figure. |

| **Supplementary Table 2.** Description of participants | | | |
| --- | --- | --- | --- |
| **Stakeholder group** | **Affiliations (non-exhaustive)** | **Sessions** | **Typical modality** |
| **Decision-makers** (national and international agencies) | **National agencies**  OGTI: General Office of Information Technologies. Ministry of Health (MINSA).  INS: National Institute of Health- MINSA  CDC-Peru: National Center for Disease Control and Prevention-Peru  DENOT: National Office for Noncommunicable Diseases, MINSA.  IETSI: Institute for Health Technology Assessment and Research-Essalud  PRONIS: National Program for Health Infrastructure Investment  Private agency: former counselor MINSA  Academia – UNMSM: Universidad Nacional Mayor de San Marcos (National University of San Marcos  Telehealth Unit - UNMSM  **International agency:**  PAHO: Pan American Health Organization | 9 interviews and 1 focus group | Virtual (video) |
| **Experts**  Professional societies & international experts | Peruvian Societies (Pediatrics, Internal Medicine, Genetics, Infectious Diseases, Hematology)  Peruvian Hereditary angioedema registry  ESID: European Societies for Immunodeficiency  NIH – National Institute of Health. USA  France (Institut Imagine) | 9 interviews | Mixed (in-person audio; virtual video) |
| **Users**  Hospital-based focus groups (Allergy and Immunology Department) | Pediatric national referral hospitals (INSN Breña, INSN San Borja)  Police and Armed Forces tertiary hospitals (National Police Hospital; Military Hospital)  EsSalud tertiary hospitals (H. Rebagliati; H. Almenara) | 1 interview and 5 focus groups | In-person (audio) |
| **Patients organizations** | Inborn errors of Immunity: “No Somos Invisibles – IDP”  Cystic fibrosis: FIQUI-Perú  Generals:  FEPER: Peruvian Federation of Rare Diseases  Los pacientes importan | 3 interviews and 1 focus group | Mixed (virtual/in-person) |

**Supplementary Table S3.** Mapping of EBP characteristics through mixed-methods triangulation

|  | Quantitative analysis | | Qualitative analysis | |
| --- | --- | --- | --- | --- |
| Data group | Accepted (≥ 75% Likert 4-5) | Review (50-74 Likert 4–5) | Suggested | Discouraged |
| General information | Sex, Registration Date,  Patient data, Informed consent, Unique patient code, Leading cause of death, Death,  Age, ICD-10, Hospital/ Institution of origin,  Identity document number, Birthdate,  Date of death, Morbidity associated with mortality, Physician in charge of the registry, Country of birth, Last name, First name, Name of contact person | Doctor's details,  Medical specialty,  Country of Origin,  Patient's phone number,  Type of kinship,  Telephone number of the contact person,  Socio-economic distribution,  Doctor's email | Unique patient identifier (national ID or registry code), date of birth/age, sex, backup contact to avoid loss of follow-up. Facility/center ID, treating service, attending clinician ID  Region/department, urban/rural, language, insurance/coverage; factors tied to access/care pathways. Selected socioeconomic indicators relevant to program goals | Full names visible in analytical views, duplicate entry  Marital status, sexual orientation, religion |
|  | Rejected (<50 Likert 4–5)  Address, Educational Level, Marital Status | |  |  |
| Clinical data | IUIS 2024 Specific Diagnosis, Genetic Diagnosis, IUIS 2024 Diagnostic Subgroup, Patient is Index Case , Date of Symptom Onset, Date of IEI Diagnosis, Affected Gene, Family History of PID, IUIS 2024 Diagnostic Group, Consanguinity of Parents, Family Diagnosis of IEI, Type of Genetic Testing Performed, IEI Relative Code, Additional Affected Genes | Twin pregnancy,  Laboratory where the genetic analysis was performed, Reason for requesting genetic analysis | Flag for suspected vs confirmed case; date of symptom onset; date of diagnosis  Principal diagnosis (ICD-10/11; ORPHANET), multiple Dx tracking; onset/phenotype tags. Mapped terminologies (LOINC/OMOP) for downstream analytics  Structured checklists (respiratory, skin, autoimmunity/inflammation, others), complications/ adverse events. Disability scales where relevant.  Key immunology labs (eg, immunoglobulins), relevant baseline values; test dates; lab facility. Lab panels tied to specific PID subtype; ability to upload reports  Genetic test performed (Y/N), test type, result status (positive/negative/variant class), gene/variant if confirmed | Recording only suspicions without minimal criteria  Free-text diagnoses; non-standard abbreviations; inconsistent coding across sites  Long open text narratives that duplicate EHR content  Unstructured lab text; over-granular panels for all patients regardless of relevance.  Detailed variant healing in every case; manual transcription of long reports |
| Therapy | Immunoglobulin Use, Gene Therapy, Registration Update Date, Transplant,  Type of transplant,  End of therapy date,  Transplanted organ,  Transplant date,  Antibiotic prophylaxis,  Start date of immunoglobulin therapy, Transplant outcome, Immunoglobulin range, Physician in charge of updating the record, Adverse effects associated with immunoglobulin,  Immunoglobulin dose,  Dosage compliance,  Date of the patient's last visit/check-up,  Reason for termination,  Last visit type/Control,  Patient weight, Route of administration,  Hospital of origin,  Patient's height, Medical specialty | Doctor's email, Immunoglobulin brand | Current/previous treatments (immunoglobulin, transplant, gene therapy, interferon- γ , anti-TNF, JAK inhibitors), start/stop dates, adverse reactions.  Visit dates, hospitalization episodes/LOS, outcomes; timeliness fields; adherence flags. Automated reminders/alerts (if available in platform) | Treatments not linked to a recorded diagnosis; free-text only without coded options.  Overly frequent forms that duplicate clinical notes; fields that can't be updated efficiently |
| Quality of life | Height, Weight  Days hospitalized, Absenteeism (in days) from school/work | Smoking | Basic, validated QoL instruments as complementary data; clearly labeled as patient-reported. Staged approach: early self-report, later clinical validation | Extensive batteries or non-validated items; using QoL to drive clinical decisions without context |

**Supplementary Table S4.** Divergent stakeholder perspectives on key technical design decisions

| Topic | Favor | ID | Against | ID |
| --- | --- | --- | --- | --- |
| Sensitive socio-demographics | “Everything is important, even religion; it all adds up to something concrete. We have to know what their reality is.” | 404-2 | “Some… doesn't seem relevant… marital status, sexual orientation, religion… shouldn't be part of the registry.” | 401-1 |
| Case status: suspected vs. confirmed | “There should be two parallels: suspicions and definitive results…” | 304-2 | “I would not… enter patients with only suspicion… you will be filled with heterogeneous data.” | 206-1 |
| Registry design | A truly complete medical history… it’s basically a detailed medical history… clinical and laboratory evaluation, treatments, procedures, adverse reactions…” | 303-2 | You’re going to create a dynamic follow-up record… very much like an electronic medical record, and they’re not the same thing.” | 104-2 |
| User only doctors | “(...) the different centers that specialize in immunology could be the only centers that are qualified or, in any case, authorized to upload information and enter new information into the registry” | 306-2 | “On the social side, I think a psychologist and a social service worker should be involved, because it's the quality of life that's affected, and not just the patient's, the family, the mother (...) To measure the economic impact on society, and finally, the cost. | 105-1 |

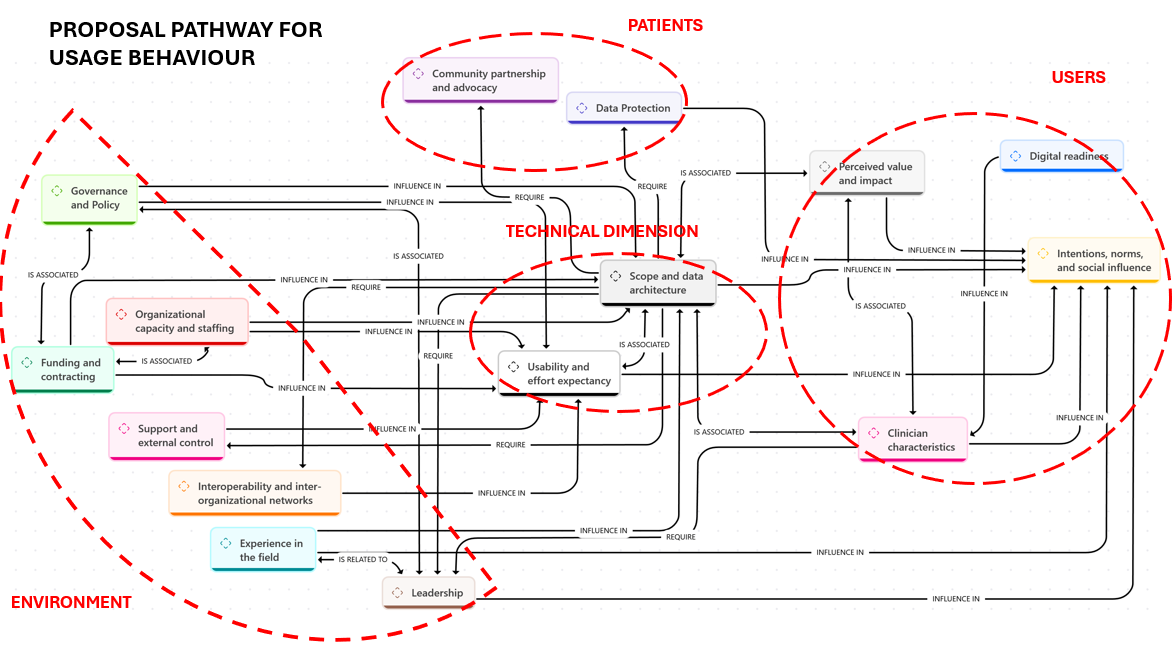

**Supplementary Figure S1. Original detailed conceptual pathway linking the four thematic dimensions and category-level relationships.** The figure summarizes the relationships identified across themes and stakeholder groups and provides a visual synthesis of how these elements were considered during the design of the registry prototype. Environmental elements such as governance, organizational capacity, leadership, funding, and interoperability are represented alongside technical aspects, including data structure and usability. User-related elements include perceived usefulness, effort, and digital readiness, while patient-related aspects include data protection and community involvement.
